# CAR-based therapeutic targets in pediatric high-grade glioma

**DOI:** 10.1101/2024.08.28.24312703

**Authors:** Myrthe M.M.R. Griffioen, Dennis S. Metselaar

## Abstract

High-grade glioma (HGG) patients have a dismal prognosis, due to a lack of effective treatments. In order to change the fate of HGG patients and decrease the current treatment-related side effects, therapy focus has shifted in the past years to immunotherapy, such as chimeric antigen receptor (CAR)-based treatments. Recent developments in CAR-based therapy show promising results in adult glioma patients, and the first clinical trials for pediatric patients with HGG are in progress. However, there are significant differences between pediatric HGG (pHGG) and their adult counterparts, including the composition of the tumor immune microenvironment (TIME), which strongly influences CAR treatment responsiveness. Therefore, we here provide a systematic overview of CAR-based therapeutic targets in pHGG entities, focusing on clinical trials and preclinical research, and comparing them to adult glioma. We conclude that target expression, TIME and CAR treatment-related toxicities vary across pHGG entities and differ from adult HGG, which suggests the need for more tailored immunotherapeutic CAR approaches in pHGG. Overall, we provide a target roadmap for future development of CAR-based therapeutic strategies for pediatric HGG patients, who are in desperate need for novel therapies.

**Graphical abstract:** 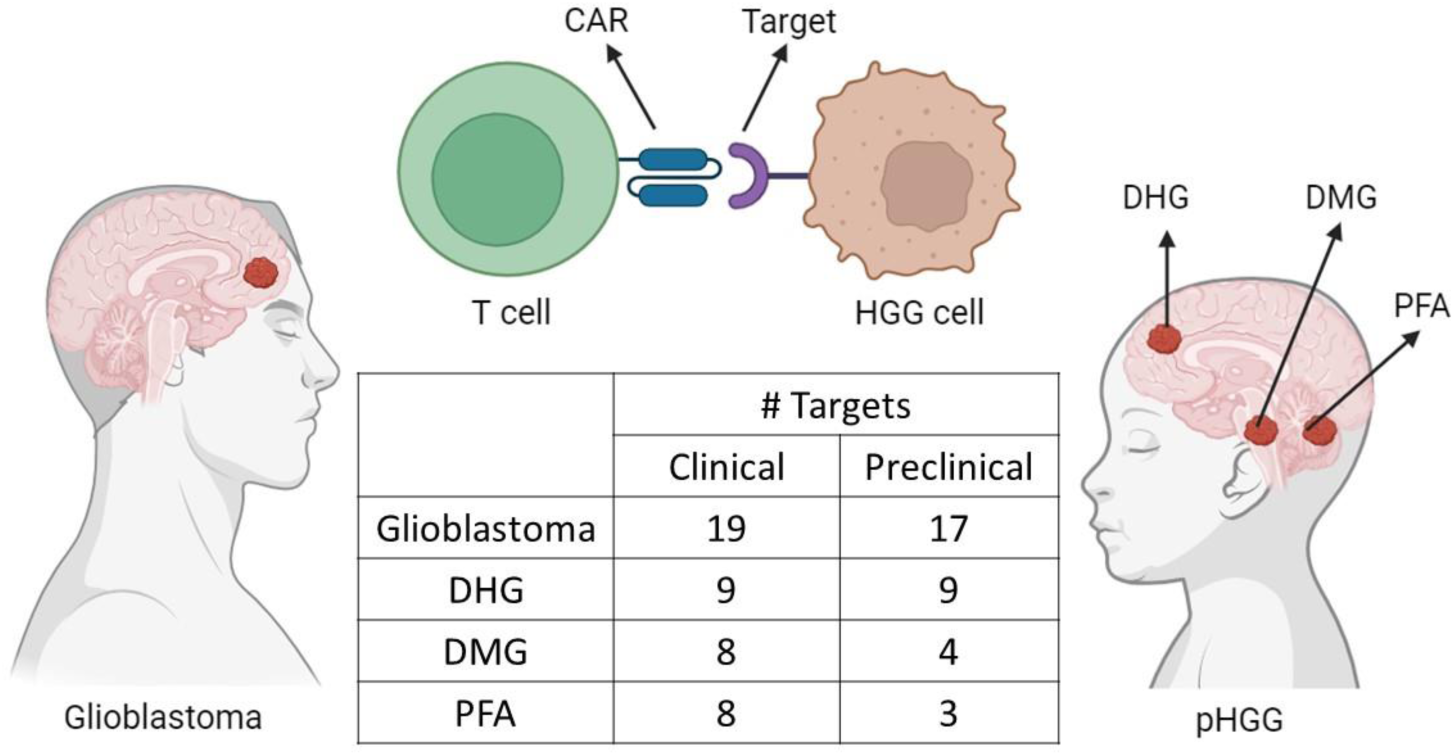

**The number of potential CAR-based therapeutic targets for glioblastoma and pHGG.** CAR: Chimeric antigen receptor; pHGG: pediatric high-grade glioma; DHG: H3 G34-mutant diffuse hemispheric glioma; DMG: H3 K27-altered diffuse midline glioma; PFA: posterior fossa ependymoma type A. Image was created in Biorender.com.

## Introduction

Gliomas arise from glial or neural stem cells and account for nearly a third of all tumors of the central nervous system (CNS) [1]. Gliomas are characterized as CNS WHO grade 1,2, 3 or 4, based on histological features and molecular markers [2]. Grade 3 or 4 gliomas are considered as high-grade gliomas (HGGs). HGGs are the most fatal group of malignancies in the CNS, with no curative options available for over 95% of its patients. Since conventional treatment options, including resection, radiotherapy and chemotherapy, often are not curative, novel therapies are needed. Recently, immunotherapies have gained significant attention as cancer treatment modality and are thoroughly investigated as potential HGG therapy. However, most clinical trials evaluating immunotherapy in HGGs only include adult patients that suffer from glioblastoma – the most common type of glioma in adults [3], IDH-mutant astrocytoma, or IDH-mutant and 1p/19q-codeleted oligodendroglioma [4].

According to the 2021 World Health Organization (WHO) classification of tumors of the CNS, pediatric gliomas are distinct from their adult counterparts in clinical presentation, gene mutations, origin, location, histological features and tumor immune microenvironment (TIME), and therefore are separately classified as pediatric-type gliomas [2]. Pediatric tumors of the CNS are the main cause of cancer-related death in children and pediatric HGGs (pHGGs) in particularly have a dismal prognosis [2, 5]. pHGGs are grouped into five different entities in the 2021 WHO classification of tumors of the CNS: H3 K27-altered diffuse midline glioma (DMG), H3 G34-mutant diffuse hemispheric glioma (DHG), H3-wildype and IDH-wildtype diffuse pediatric-type high-grade glioma (WT-DHG), infant-type hemispheric glioma (IHG) and pediatric ependymomas.

The first group, DMG, including pontine tumors in the pons, previously referred to as diffuse intrinsic pontine glioma (DIPG), are characterized by an alteration in histone 3 (H3) lysine 27 (K27). Those tumors originate in the midline of the brain, have an infiltrative nature and a median survival of 9-11 months post-diagnosis [6]. The delicate location of DMGs, and DIPGs in particular makes complete resection impossible. Radiotherapy, the standard treatment for this patient group lengthens survival with a few months [7]. The second group of pHGGs, K34-altered DHG, is characterized by an alteration in histone 3 (H3) glycine 34 (G34). Those tumors can arise anywhere in the supratentorium and have a median survival of 9-15 months post-diagnosis [8]. The third group, WT-DHG, entails a heterogeneous group of rare HGGs without H3 or IDH mutations. This entity is only recently introduced and clinical data is scarce [9]. The fourth group, IHG, occurs in infants and newborns with mutations in other genes than *H3* or *IDH*. These rare tumors can be subdivided into three groups: pHGGs that are located in the cerebral hemispheres with alterations in one of the genes *ALK*, *ROS1*, *NTRK1/2/3* or *MET*; pHGGs that are located in the hemispheres with RAS/MAPK pathway alterations; or pHGGs that are located in the midline structures with RAS/MAPK pathway alterations.

The fifth group, pediatric ependymomas (EPNs), were only recently classified under the gliomas, since they originate from radial glial cells, instead of ependymal cells, which was previously assumed. EPNs are found along the entire neuroaxis in both adults and children. However, most EPNs in adults are located in the spinal cord, accounting for ∼5% of adult CNS tumors, while pediatric EPNs mainly occur intracranially (both supratentorial and infratentorial) accounting for ∼10% of all CNS tumors [10]. Based on molecular profiling, nine different EPN subtypes can be distinguished with different clinical, (epi)genetic and prognostic properties. Besides three groups of spinal (SP) EPNs, two main types of supratentorial (ST) EPNs are found in children, both characterized by gene fusions (ST-EPN-ZFTA and ST-EPN-YAP), and two types of posterior fossa (PF) EPNs: PF-EPN-A (PFA) and PF-EPN-B (PFB). Of those, PFA, is the most common and most fatal ependymoma [11]. PFA EPNs are characterized by a loss of H3K27trimethylation, which is a shared hallmark with DMGs. Treatment of EPNs entails resection, followed by radiotherapy and in a few cases chemotherapy [12].

Besides the calamitous mortality rate of HGG patients, most survivors face devastating treatment-related co-morbidities. These adverse effects can be especially serious for pediatric patients, whose treatments affect the developing brain [13]. Therefore, there is an urgent need for novel tumor-specific targeted therapies such as immunotherapies that do not harm surrounding normal brain tissues. In recent years, immunotherapy has gained a strong momentum as potential cancer therapy, of which chimeric antigen receptor (CAR) T-cell therapy is one of the most explored ones. CAR T cell-based therapy relies on the manufacturing of modified patient-derived or off-the-shelf T cells that display a chimeric antigen receptor that specifically binds to a tumor associated antigen (TAA) [14]. After binding of the receptor to the TAA, the T cell gets activated and differentiates to either a CD4+ T cell that releases cytokines for activation of the immune system or a CD8+ T cell that destroys the cancer cell. Currently, there are several FDA-approved CAR-based therapies for B cell malignancies, like anti-CD19 CAR T cells and anti-BCMA CAR T cells [15].

Currently, many clinical trials are evaluating CAR T cell therapy for other malignancies such as HGG as well. Despite some challenges including heterogeneous expression of TAAs, route of administration, immunosuppressive TIME and treatment-related toxicity, clinical reports of CAR T cell trials in HGGs are promising. Most studies on CAR-based therapy for gliomas focus on adult glioblastoma. Since childhood HGGs are characterized by different mutations and show distinct TAAs and TIME compared to their adult counterparts, more tailored CAR target selection will be crucial for future treatment of pHGGs. Currently, the first clinical trials focusing on CAR-based therapy for pHGG and a wide-variety of preclinical studies in these malignancies are ongoing. In this review, we summarize the targets and strategies for CAR-based therapy for pHGG and compare them with targets described for adult glioblastoma. We evaluate their status, future prospects, and discuss why distinct CAR-based therapy targets and strategies are needed for pHGG.

## CAR targets in HGG

We collected data from clinical trials on CAR-based therapies for HGG, including age of the patients, trial phase, CAR-based target and glioma entity (**Supplementary table 1**). To date, most completed and ongoing clinical trials evaluating CAR-based therapy in glioma only included adult patients (**Figure 1**). The top five antigens in clinical trials for HGG are: EGFRvIII, B7-H3, IL13Rα2, HER2 and GD2. Although research describing CAR-based therapeutic targets in pHGG is sparse compared to adult glioblastoma, there are currently eighteen ongoing clinical trials in pHGG, with some preliminary data reports (**Supplementary table 1**). We will now further elaborate the results that each of these CAR targets has yielded in pHGG.

**Figure 1.**
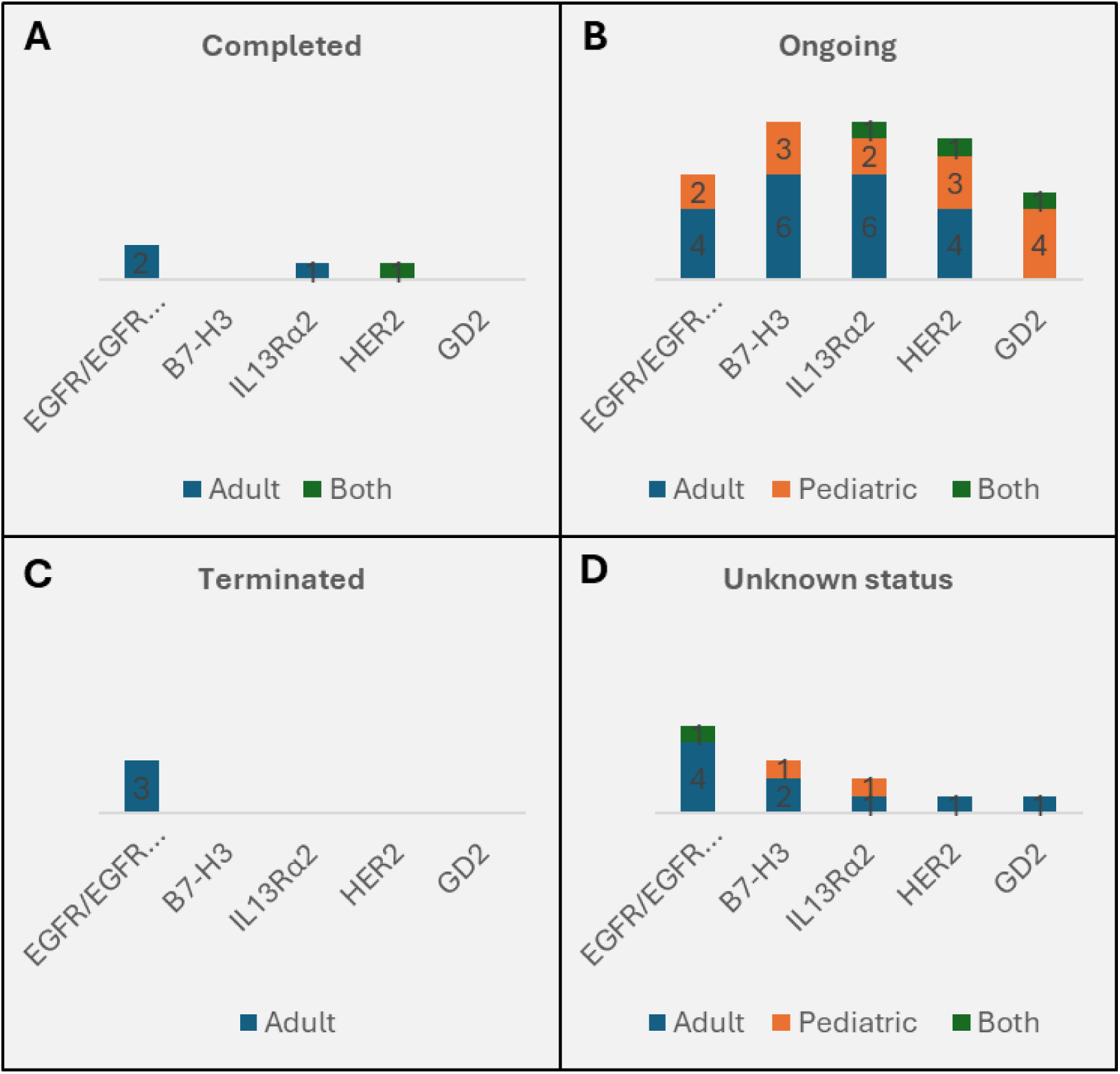
Bar graphs showing the number of clinical trials for the top five CAR-based therapeutic targets in HGGs. Clinicaltrails.gov was accessed on 15-01-2024. (**a**) Completed clinical trials; (**b**) Ongoing clinical trials; (**c**) Terminated clinical trials. (**d**) Clinical trials with an unknown status. View details of all clinical trials in **Supplementary table 1.**

### 2.1. EGFR/EGFRvIII

To date, most clinical trials on CAR-based therapy for HGG used anti-EGFRvIII CARs. The transmembrane protein EGFR (ERBB-1) is a member of the epidermal growth factor (EGF) family. EGFR plays a role in the migration of neural stem cells during development and activation of the MAPK and PI3K-Akt pathways to promote cellular proliferation. The EGFRvIII variant arises from a deletion of exon 2-7 in the *EGFR* gene, which results in a functional protein with a mutation in the extracellular domain. The *EGFR* gene is expressed in normal tissue, but is amplified in 30-40% of glioblastoma, with overexpression in ∼75% of the cells [16–18]. The EGFRvIII variant is specific for adult glioblastoma (present in ∼30% of patients with ∼75% EGFRvIII-positive cells). [19–22]. Several preclinical studies in murine models found that anti-EGFRvIII CAR T cells were effective [23, 24].

O’Rourke et al. (2017), reported the first 10 adult glioblastoma patients that received a single dose of intravenous infusion of anti-EGFRvIII CAR T cells and found that this resulted in on-target trafficking of CAR T cells to the tumor (NCT02209376) [25]. One patient had extended survival, although treatment resulted in antigen loss and increased immunosuppressive inhibitory molecules and regulatory T cells in the TIME [25, 26]. This study also reported that PD1 expression in the CAR T cell product positively correlated with engraftment in peripheral circulation and progression-free survival (PFS), which contradicts earlier research that characterized PD1 as a marker of T cell exhaustion [27]. In fact, several clinical trials focused on blocking the immune checkpoint PD1 with antibodies to improve T cell activation [28]. Furthermore, a combinational therapy with anti-EGFRvIII and PD-1 inhibitors in a murine glioblastoma model was found to be effective [29]. However, infusion of anti-EGFRvIII CAR T cells in combination with an anti-PD1 antibody was not effective in patients (NCT03726515; [30, 31].

A dose escalation study from Goff et al. (2019) also did not find clinical responses to the anti-EGFRvIII CAR in 18 glioblastoma patients in an altered protocol with another design (NCT01454596) [32]. Patients that received the highest treatment dose showed dyspnea/hypoxia, a dose-limiting pulmonary toxicity, which was fatal for a patient that received the highest dose. The authors suggest that delivery of the CAR T cells intrathecal, intratumoral or intraventricular, instead of intravenous, could improve the efficacy against glioblastoma. To conclude, currently, none of the clinical studies on EGFRvIII-targeting CAR T cells observed regression of the glioblastoma or other treatment-related clinical benefits.

Besides the presence of the EGFRvIII variant, many adult gliomas also overexpress wild-type (WT) EGFR, which could be a potent target for CAR-based therapy as well. However, since the expression of EGFR is not limited to cancer cells, CAR therapy targeting WT EGFR may result in off-target toxicities in astrocytes or keratinocytes. Therefore, a novel CAR design was recently developed that specifically targets amplified EGFR, EGFRvIII and other EGFR extracellular domains with mutations [33]. This CAR (806EGFR) does not recognize wild-type EGFR on normal brain tissue, since the epitope is only exposed in conformations that are present on overexpressing tumor cells [34]. The approach to use a CAR with broad specificity to EGFR alterations could have the advantage of targeting multiple clones in the tumor and reducing the risk of antigen loss, as was seen in clinical studies that selectively targeted the EGFRvIII variant (NCT02209376). No clinical reports are yet published on the use of 806EGFR CAR T cells for glioblastoma, although it was shown to be effective in preclinical studies in glioblastoma [35, 36]. In addition, several preclinical studies also showed effectivity of a dual EGFR/EGFRvIII CAR approach [37, 38]. Other novel anti-EGFRvIII CAR designs are currently tested in preclinical research, such as an optimized anti-EGFRvIII CAR that showed effectivity in orthotopic models of human glioblastoma in NSG mice [39].

EGFR amplification is found in 11% of pHGGs, 3% of pediatric ependymomas, and 8.6% of the pHGGs that show EGFR amplification harbored the EGFRvIII mutation [40, 41]. Given the fact that EGFRvIII deletions are not as common in pHGGs compared to glioblastoma, and the disappointing clinical results from EGFRvIII CAR T cells targeted at glioblastoma in patients, EGFRvIII might not be a promising target for CAR-based therapy in pHGGs. Still, there are currently two clinical trials on EGFR/EGFRvIII for pHGGs, without any clinical data reported yet. Clinical trial NCT03638167 investigates whether CAR targeting the EGFR806 epitope is effective for pediatric EGFR-positive recurrent or refractory CNS tumors (excluding DMG). Clinical trial NCT05768880 investigates the use of a quad CAR targeting four antigens at once, including EGFR/EGFRvIII in pediatric recurrent or refractory CNS tumors.

### 2.2. B7-H3

The second most investigated target in clinical trials on CAR-based therapy for HGG is B7-H3 (CD276). The transmembrane protein B7-H3 is a member of the B7 protein family, and plays a role in inhibition of the immune response. B7-H3 mRNA is widely expressed in normal and cancer tissue, but while normal tissue has limited expression, the protein is overexpressed in multiple human cancers, including glioma [42]. Around 70% of adult glioblastoma shows overexpression of B7-H3, with 95% of the cells expressing B7-H3, correlating to a worse prognosis [43–45]. In children, B7H3 is highly expressed in pHGG, with ∼75% of non-DMG showing B7H3 expression and all tested DMG being-positive with a tumor cell percentage of ∼90%, reducing the risk for antigen loss [46–48]. Several preclinical studies found efficacy of anti-B7-H3 CAR T cells in mouse models with xenograft glioblastoma or glioma [44, 45, 49, 50]. Haydar et al. (2021) showed inhibition of DMG progression in patient-derived orthotopic xenograft (PDOX) models and increased survival after administration of CAR B7-H3 T cells [47].

In addition, Majzner et al., 2019 tested B7-H3 CAR T cells in xenograft mouse models of several pediatric solid tumors and showed antitumor activity, dependent on the antigen density on tumors, which resulted in sparing the normal tissues that have very low expression of B7-H3 [48]. Preliminary results from Mehta et al. (2023) show that anti-B7-H3 CAR T cells also had antitumor activity in a ST-EPN-ZFTA xenograft mouse model, but that tumors recurred [51].

Preliminary data from the first clinical trial that reported a case report on an adult patient with glioblastoma receiving several intracavitary infusions of anti-B7-H3 CAR T cells showed that there was a clinical response (reduction of the recurrent glioblastoma) [52] (ChiCTR1900023435). However, the tumor recurred after cycle 6-7 and patient experienced drowsiness and altered consciousness. The main side effect was headache. After 7 cycles, the patient decided to resign from the clinical trial. This clinical trial, among six other clinical trials are currently ongoing testing anti-B7-H3 CAR T cells as a treatment for glioblastoma.

Given the promising results from preclinical studies and the fact that B7-H3 is highly expressed on pHGGs, especially in DMGs, there are currently several phase 1 clinical trials exploring the locoregional or intravenous delivery of anti-B7-H3 CAR T cells for the treatment of pHGGs. One of them reported preliminary clinical findings of intraventricular delivery of B7-H3 CAR T cells in two patients with DMG and one patient with IDH-mutant astrocytoma [50] (NCT04185038). Although the B7-H3 expression was not evaluated, the DMG patients showed some increased survival, albeit with progressive tumor growth. The astrocytoma patient showed stable disease, and even a decrease in tumor size of 19%, but discontinued the study after 1 year. The main side effects were headache, nausea and fever. In a subsequent trial update 13 DMG patients were reported to have been treated without occurrence of cytokine release syndrome (CRS) or immune effector cell-associated neurotoxicity syndrome (ICANS), although one patient had dose limiting toxicity (pontine hemorrhage, eight days after the initial dose). In addition, analysis of the cerebrospinal fluid (CSF) showed presence of CAR T cells and immune activation [53].

### 2.3. IL13Rα2

The third most investigated target in clinical trials on CAR-based therapy for HGG is IL-13Rα2 (CD213A2), a transmembrane protein and subunit of the IL-13 receptor complex. IL-13Rα2 regulates immune response by binding to cytokine IL-13. IL-13Rα2 is overexpressed in several cancers, including pancreatic cancer, breast carcinoma and ovarian cancer, and not expressed in normal tissues except for the pituitary and testis [54]. IL-13Rα2 is overexpressed in around 50% of glioblastoma and 85% of DMGs, and high expression correlates to poor prognosis [55]. IL13Rα2 is overexpressed in 65-80% of pediatric ependymomas, with 55% positive cells [47, 56]. Several preclinical studies in murine models found that anti-IL13Rα2 CAR T cells were effective against glioblastoma and DMG [57–59].

Brown et al. (2015) reported a phase 1 safety and feasibility report of three patients with recurrent glioblastoma who received up to twelve cycles of intracavitary or intratumoral infusions of anti-IL13Rα2 CAR T cells (NCT00730613) [61]. The main side effects were neurological deficits and headache. Two patients demonstrated an anti-tumor response which was characterized by tumor necrosis and presence of T cells in the tumor. However, there was a reduction in IL13Rα2 expression in the tumor cells (antigen loss), and tumor cells with low IL13Rα2 expression continued to proliferate. All patients had tumor recurrence which resulted in death.

A dose escalation study by the same author demonstrated tumor regression in one patient which lasted for 7.5 months after intracavitary and intraventricular infusions (NCT02208362) [62], with intraventricular infusion outperforming the intracavitary administration. In 2024, the same group reported results from the completed phase 1 trial on 65 patients (12-75 years old) with recurrent HGG that were treated with weekly intratumoral and/or intraventricular infusions of anti-IL13Rα2 CAR T cells [63]. They described that repetitive locoregional infusions of anti-IL13Rα2 CAR T cells are feasible, there were no dose-limiting toxicities (DLTs) reported and the most common side effects were headache, fatigue and hypertension.

Wang et al. (2023) evaluated the treatment with anti-IL13α2 CAR T cells in six patients with pHGG and reported radiographic benefit in two DMG patients, who both achieved stable disease (NCT04510051) [64]. Two PFA ependymoma patients showed no response and the tumors had low levels of T cell infiltration and high levels of myeloid infiltration. One PFA ependymoma patient had a reduction in tumor size of the largest tumor, however, he also had progression of multifocal and leptomeningeal tumors. All patients had an increased proportion of CAR-negative T cells in the CSF over time.

### 2.4. HER2

The fourth most investigated target in clinical trials on CAR-based therapy for HGG is HER2 (CD340 or Neu or ERBB-2). The transmembrane protein HER2 is a member of the epidermal growth factor (EGF) family, belonging to the same family as EGFR. HER2 also plays a role in cellular proliferation and its expression is correlated to poor prognosis in glioblastoma [65]. HER2 is overexpressed in several cancers, including glioma and shows low expression in normal tissues [66, 67]. HER2 is overexpressed in 30% of glioblastoma and 60-65% of cells is HER2-positive [18, 68]. 40% of DMG, 40% of pediatric ependymomas and 75% of other pHGG is HER2 positive, with 35-65% HER2-positive cells [47]. Preclinical research in murine models found that anti-HER2 CAR T cells were effective against glioblastoma and DMG [69, 70].

Ahmed et al. (2017) reported results on a first phase 1 safety, feasibility and dose-escalating clinical trial testing intravenous infusion of anti-HER2 CAR T cells as a treatment for HGGs in 10 adults and seven children (<18 years old) (NCT01109095) [71]. They found that the treatment was safe and found no dose-limiting toxicity. Moreover, they showed clinical response of reduction in the tumor size and extended survival in one of the adolescent patients and stable disease was achieved in seven patients.

Preliminary results of a clinical trial testing repeated locoregional infusion of anti-HER2 CAR T cells as a treatment for pediatric CNS tumors showed that the CAR treatment is safe and induces an immune response (NCT03500991) [72]. In addition, there is a clinical trial is ongoing testing intravenous infusion of anti-HER2 CAR T cells as a treatment for pediatric ependymomas (NCT04903080).

### 2.5. GD2

The fifth most investigated target in clinical trials on CAR-based therapy for HGG is GD2. The ganglioside GD2 is a member of the glycosphingolipid family [73]. Its cancer-associated role is to promote cell survival, invasion and T cell dysfunction, as well as functioning as an immune checkpoint. GD2 is widely expressed on several solid tumors, with a special focus on GD2 as a target in pediatric neuroblastoma, where anti-GD2 CAR T cell therapy is promising [74, 75]. GD2 expression is found in 100% of pHGGs with 80-90% GD2-positive cells; and 40% of pediatric ependymomas with 80% GD2-positive cells [47]. In pHGG, including DMG, a heterogeneous GD2 expression has been reported in patient-derived samples, with low expression in non-tumor CNS tissue [77]. However, heterogeneity is lost in DMG cell cultures, resulting in a high, homogenous expression in DMG and low levels in H3-WT cells [76, 77]. Preclinical research in murine models found efficacy of anti-GD2 CAR T cells against both adult glioblastoma and pHGG [76–80].

Majzner et al., 2022 published the results of the first four patients with DMG who received several intravenous infusions with anti-GD2 CAR T cells, and one patient receiving intracerebroventricular infusions (NCT04196413) [81]. This study showed that the patients experienced cytokine release syndrome (CRS) and tumor inflammation-associated neurotoxicity (TIAN) that had to be treated on the intensive care unit, where drainage of CSF was necessary. However, treatment also resulted in a reduction in neurological symptoms and radiographic improvements. In one of the patients with spinal cord DMG, a reduction of 90% of the tumor volume was observed after the first infusion. Another patient showed infiltration of T cells in the tumor, but without clinical benefit. A third patient had 27% reduction of tumor volume after five infusions, however he died due to intratumoral hemorrhage, a complication that is seen more often in DMG patients.

Liu et al., 2023 reported results on four pediatric and four adult glioblastoma patients, and showed that intravenous infusions (in three cases followed by intracavitary infusions) of anti-GD2 CAR T cells was safe (NCT03170141) [82]. No response was reported in two patients and a partial response in the other two patients. Moreover, there was evidence for antigen loss and increased infiltration of T cells and macrophages in the tumor in one of the glioblastoma patients.

In addition, a preliminary report was published by Lin et al., 2023 evaluating intravenous anti-GD2 CAR T cell infusions in pediatric CNS tumors (NCT04099797) [80]. Here, cohorts that received anti-GD2 CAR T cells with co-expression of a constitutively active IL-7R were included. Six patients were treated with the CARs with co-expression of IL-7R, and patients experienced neurotoxicity and cytokine release syndrome (CRS). Although the clinical results were promising, further analyses with larger cohorts are needed to improve the clinical benefits of the treatment.

### 2.6. Other potential targets for pHGGs

Another target that is getting a lot of focus in preclinical and clinical CAR-based research for HGG is EphA2. The transmembrane protein EphA2 is an Eph receptor linked to carcinogenesis and tumor progression in several cancers including glioblastoma [83]. Preclinical studies evaluating anti-EphA2 CAR T cells in a murine model with glioblastoma showed promising therapy efficacy [84]. Preliminary reports on EphA2 [85] showed that intravenous infusions of anti-EphA2 in three glioblastoma patients resulted in transient clinical efficacy. However two of the three patients experienced lung edema, which can be explained by the expression of EphA2 in normal proliferating epithelial cells [83, 86]. Due to these disappointing results and the lower expression levels of EphA2 in pHGG, EphA2 might not be a promising target to investigate for pediatric glioma.

Currently, several promising targets have been investigated in preclinical research, mostly focusing on glioblastoma: αvβ3, PDPN, CAIX, GRP78, CSPG4, FAP, Fn14, CD317, EphA3 and P32 [87–95], and many (adult only) glioblastoma clinical trials testing other targets are ongoing: CD133, CD147, CD44, CD70, EphA2, MMP2/Chlorotoxin, MUC1, NKG2D ligands and PD-L1 (**Supplementary table 1**). However, only two ongoing clinical trials are testing potential targets in pHGG; one trial is exploring a treatment with a CAR targeting WT1, PRAME and Survivin in adult and pediatric HGG (NCT03652545) [100–102], based on results from studies in the leukemia field. Another trial is assessing a CAR targeting H3.3K27M as a treatment for DMG (NCT05478837); although some preclinical data suggests that a CAR targeting an HLA-A∗02:01 restricted histone 3K27M epitope in DMG is not effective because of lack of presentation [98, 99], this trial is based on some promising preclinical results that include *in vivo* data [96, 97].

Recent preclinical studies found efficacy of CAR T cells in a murine pHGG model, targeting other TAAs such as integrin αvβ3, GPC2, CD99 and PDGFRα [87, 103–105]. Other potential targets were found in studies aiming to find immunotherapy targets by analyzing the upregulation of membrane proteins in glioma tissue/patient-derived cell lines compared to normal tissues. Here, gp100, MAGE-1, TROP2 (in glioblastoma), FAP and TNC (in both glioblastoma and pHGG), OAcGD2 (in pHGG), EphA3, GRP78 and neogenin (in DMG) were identified [106–113].

### 2.7. Multi-target CAR approach

Since TAAs are often heterogeneously expressed in gliomas, conventional mono-target CAR T cell strategies are often insufficient due to antigen loss. Therefore, several preclinical and clinical studies are now evaluating a dual-tri-or quad-CAR approach. A dual CAR approach, where CAR T cells express two distinct CARs against two different antigens, is currently under investigation for glioblastoma in three phase I clinical trials (NCT05168423; NCT05577091; NCT05868083). In addition, tandem CAR T cell strategies, where one CAR molecule targets two different antigens, showed promising preclinical efficacy in a murine glioblastoma model [114–116].

There is one clinical trial investigating Tri CARs targeting WT1, PRAME and Survivin in adult and pediatric recurrent and refractory CNS tumors (NCT03652545). A CAR T cell with three distinct CAR molecules targeting HER2, IL13Rα2 and EphA2 showed promising results in glioblastoma and PFA ependymoma murine models [68, 117]. Currently, a phase I clinical trial of repeated locoregional infusions of quad CAR T cells against EGFR/EGFRvIII, HER2 and IL13Rα2 for children with recurrent/refractory CNS tumors and DMG is ongoing (NCT05768880). In addition, there are preclinical promising results of T cells with CARs against HER2, EGFR806, B7-H3 and IL13Rα2 in glioblastoma and DMG murine model [118]. Novel multi-target strategies also involve co-expression in CAR T cells of receptors to bind to TAAs [119]. Although preclinical results are promising, no clinical reports are yet published on the use of multi-targeting CAR T cells in glioblastoma or pHGG patients.

## Discussion

In this review, we explored the literature on CAR-based therapy targets for pHGG and glioblastoma. Most CAR-T clinical trials focus on adult glioblastoma, and most antigens that were proposed for pHGG, such as EGFRvIII, HER2 and IL13Rα2, derive from these studies. However, expression of these antigens is often much lower in pHGG compared to adult glioblastoma, indicating that targeting these antigens will likely be less effective in pHGG. In addition, we found that the expression levels of abovementioned TAA vary across the different entities of pHGG (**Supplementary table 2**), indicating that a tumor-entity specific approach is required.

**Figure 2.**
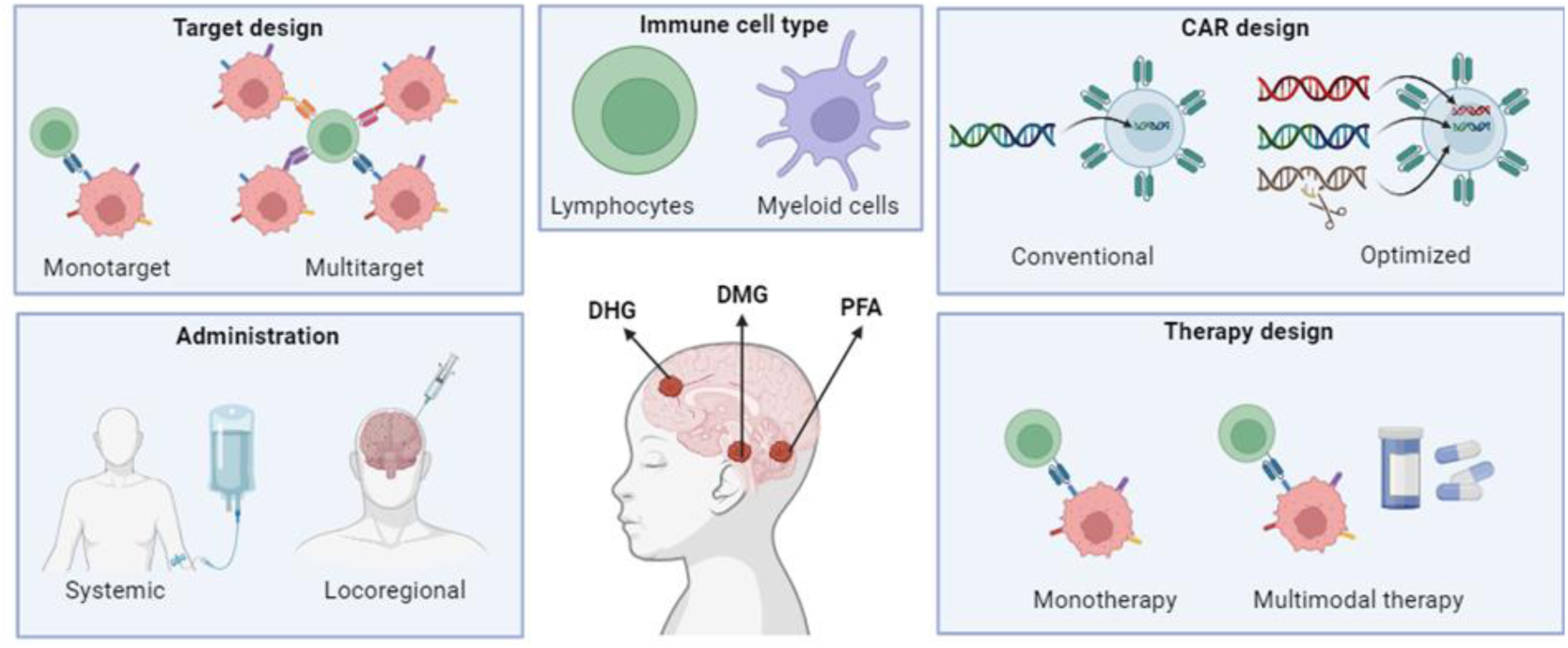
Overview of CAR-based therapy optimizations. DHG: H3 G34-mutant diffuse hemispheric glioma; DMG: H3 K27-altered diffuse midline glioma; PFA: posterior fossa ependymoma type A. Image is created with Biorender.com.

### 3.1. Novel CAR T cell designs

Several clinical reports on adult and pediatric HGG targeting a single TAA that is not uniformly expressed report loss of the antigen [25, 26, 61, 82]. Due to this antigen loss, the antitumor effect of CAR T cells is reduced, resulting in treatment resistance and tumor recurrence. This is alarming for adult glioblastoma and pHGGs, where uniformly expressed TAAs are lacking, and normal tissue often also shows expression of the antigen. Both adult and pediatric HGGs are extremely heterogeneous and consist of distinct subpopulations of glioma cells, with variable antigen expression. Therefore, targeting multiple TAAs could be a strategy to overcome antigen loss. Targeting TAAs that also show low expression in normal tissues, or targeting multiple TAAs increases the risk of adverse effects due to on-target off-tumor toxicity. However, these side effects might not outweigh the clinical advantages of multi-targeting for pHGG patients, since no other therapy is available for pHGG.

In addition, logic-gating CAR T cell designs which increase tumor cell targeting or reduce off-target risk have been proposed, such as Tandem, SynNotch and REV CARs [120]. Tandem CARs recognize two antigens with a single CAR molecule, which reduces antigen escape [114]. SynNotch CAR is a design where the activation of one CAR molecule – after binding to its antigen -induces the transcription of another CAR molecule specific for a second antigen, therefore avoiding off-target activity and allowing increased tumor specificity [121]. Switchable REV CARs have a peptide epitope as their extracellular domain and a bispecific target module that recognizes the epitope and a TAA which activates the CAR T cells, allowing for increased tumor specificity [122]. These are examples of future CARs that will enhance tumor specificity and reduce side effects, which is particularly relevant for pHGG patients.

For optimal anti-tumor efficacy, CAR designs need to be optimized to improve effective antigen binding domains and signaling domains. CAR designs have been increasingly optimized over the past years, giving rise to a variety of CAR generations [123]. Initially, CAR T cells were thought to require high antigen expression on tumor cells for effectiveness [124]. However, several preclinical studies demonstrated that precise CAR design could influence the threshold for antigen recognition [112, 125–127]. Another approach for improved tumor targeting is the use of bispecific T cell engagers (BiTEs) or bispecific killer cell engagers (BiKEs), which are secreted by the T or NK cells [128, 129]. Currently, one clinical trial is combining dual anti-IL13Rα2 and anti-HER2 CAR T cells with EGFR and EGFRvIII-specific BiTEs in glioblastoma patients (NCT05868083). These novel CAR designs with improved tumor targeting, implicate that pHGG antigens that were initially not considered potential TAAs for CAR-based therapy due to their relatively low expression, can now be reconsidered and tested in preclinical and clinical studies.

### 3.2. Role of the TIME in CAR T cell therapy

CAR T cell treatment is often hampered by the immunosuppressive TIME of these malignancies, which limits the infiltration of the CAR T cells into the tumor [130–133]. In fact, it has been reported that WT-DHG has an anti-inflammatory microenvironment, and DMG has a dampened or non-inflammatory microenvironment [133]. In both WT-DHG and DMG, there is high infiltration of M2-like myeloid cells and limited infiltration of T cells [133]. In contrast, PFA has a pro-inflammatory microenvironment, with high infiltration of both M1-like myeloid cells and T cells [133, 134]. However, Wang et al. (2023) reported that anti-IL13Rα2 CAR T cells were ineffective in two PFA ependymoma patients, whose tumors had low levels of T cell infiltration and high levels of myeloid infiltration [64]. Therefore, it is likely that modulation towards a pro-inflammatory TIME with high T cell infiltration is required for effective CAR treatment in pHGG. This could have a considerable influence on the way we develop immunotherapies, because we need to develop synergistic therapies, which modulate the TIME in parallel to targeting the glioma cells by CAR T cells.

Haydar et al. (2023) found that inflammatory myeloid cells are essential of effective CAR treatment [135]. In addition, they found that treatment with CD28-based CARs resulted in upregulation of genes associated with inflammatory myeloid responses, whereas treatment with CARs with other signaling domains resulted in upregulation of genes associated with suppressive myeloid responses and dysfunctional T cell responses. This result indicates that CAR-based therapies for pHGG could benefit from using tumor-entity specific CAR designs that are most effective in the tumor-entity specific TIME.

Recently, several studies found promising preclinical results of co-expression or blocking of immunomodulatory molecules in the CAR T or NK cells [79, 136–139]. This is relevant to consider for developments of novel CAR T cell therapies for pHGG with anti-inflammatory or non-inflammatory TIME, since the expression or knock-out of immunomodulatory proteins can lead to simultaneous targeting of the TIME, or even modulate the TIME towards a more favorable pro-inflammatory TIME. Another novel strategy to overcome the negative influence of the TIME and improve CAR cell infiltration in pHGG is to target not only the glioma cells, but also the vasculature or microenvironment of the tumor, and has been explored in several preclinical studies [93, 140, 141].

### 3.3. Route of administration

Standardly, clinical trials delivered the CAR T cells via intravenous infusions, which is the least invasive method. However, several novel studies now indicate that locoregional infusions, such as intraventricular, intrathecal, intracavitary and intratumoral administration, are more effective than intravenous infusions [62, 81, 117, 142, 143]. The authors in these studies claim that locoregional administration is the preferred route of administration, because it enhances CAR T cell trafficking to the glioma cells and requires lower dosage than systemic delivery, which reduces the on-target off-tumor risk. Also, in DMG specifically, the blood brain barrier (BBB) is relatively intact, which might hamper the trafficking of CAR T cells towards the glioma [144]. However, even with locoregional administration, there is still no guarantee that the CAR T cells will spread throughout the whole tumor and also towards the diffuse tumor cells, which are located at a larger distance from the infusion site. In addition, to this date, convincing clinical evidence from large cohorts comparing systemic and locoregional CAR T cell infusions is lacking.

### 3.4. Treatment-related toxicity

Majzner et al. (2022) showed that locoregional administration of anti-GD2 CAR T cells in DMG patients resulted in severe side effects, they indicated as TIAN [81]. This CAR T cell treatment-related swelling in the brainstem is extremely dangerous, and can be fatal. Therefore, preclinical studies focus on novel strategies to improve CAR T cell treatment safety in DMG to reduce toxicity. For example, intratumoral infusion of mRNA CARs are under development for a safer treatment in DMG [145]. Still, the severe side effects raise the concern that immunotherapy might not be feasible or will result in long-term adverse effects, especially in pediatric patients whose brains are still developing. Therefore, since it is likely that CAR treatments will result in higher survival rates of pHGG patients in the future, CAR treatments will also create new socio-economic problems, such as loss of motor and cognitive functions in cured patients, and a higher burden on healthcare.

### 3.5. Novel CAR-based cell types

Recently, several preclinical studies found promising results of using γδ T, NK and myeloid CAR cells, instead of conventional CAR αβ T cells in targeting of adult and pediatric HGG [110, 136, 146–151]. The main advantage of γδ T cells is their higher antigen recognition options, for instance by also being able to target soluble proteins in the TIME [152]. This can be of potential for CAR treatment of pHGG, because optimal TAAs for CAR T cell therapy in pHGGs are currently lacking and novel therapeutic targets are required. Therefore, it is likely that broadening the antigen recognition by including soluble proteins as well, will increase CAR-based therapeutic options.

The main advantage of myeloid cells, such as macrophages, microglia and neutrophils, is the observation that the proportion of myeloid cells in pHGG is much higher than the proportion of T cells [133, 153], suggesting that myeloid CARs may have a significantly improved infiltration and anti-tumor efficacy in pHGG. The main advantage of NK cells, is that administration of allogenic NK cells is safe, whereas allogenic T cells can induce graft-versus-host disease [154]. This will reduce costs and production time of development of CAR T cell treatments. The reduction of waiting time before the start of the CAR T cell treatment is especially important for pHGG patients, because of their short survival after diagnosis.

### 3.6. Combination therapies

Although CAR-based immunotherapies are promising, there are some challenges regarding tumor infiltration and immune activation, which potentially require multimodal treatment strategies. Therefore, several studies found that combination therapy with chemotherapy or radiotherapy can enhance the efficacy of CAR T cell therapy. The use of lymphodepleting chemotherapy is already common procedure in CAR therapy [155]. It is shown in hematological diseases that lymphodepletion prior to CAR T cell therapy modulates the immune environment, characterized by reduction of immunoregulatory cells, which is beneficial for the efficacy of the treatment. Several preclinical studies now indicate that combination therapy of CAR T cells and chemotherapy or radiotherapy also results in synergistic efficacy in adult and pediatric HGG [77, 155–160]. However, currently there is no data validating its clinical efficacy in peripheral or locoregional administration of CAR-based therapies for brain tumors. Thus, although lymphodepleting chemotherapy prior to CAR T cell therapy is expected to modulate the TIME and enhance CAR T cell activity in pHGG, this needs to be confirmed in clinical trials.

Since it has been reported that radiotherapy can increase the expression of stem-cell like markers CD133, CD70 and EphA2 in glioblastoma, it has been proposed as a combination therapy in stem cell marker-targeting CAR therapies [160]. There is currently no data on the effect of radiotherapy on stem-cell like marker expression in pHGG. However, since these stem-cell like markers are relatively low expressed in pHGG, this might not be as effective for pHGG as for glioblastoma.

Standardly, corticosteroids such as dexamethasone, are used in DMG treatment to prevent fluid accumulation in the brain. Since adverse effects, such as swelling of the brain, were reported by Majzner et al. (2022) as a result of CAR T cell treatment in DMG patients [81], CAR treatment could be combined with these corticosteroids in the future. Moreover, recently a preclinical study in an ovarian tumor murine model found that dexamethasone enhances CAR T cell persistence [161]. These data suggest that dexamethasone could enhance CAR T cell treatment in pHGG patients, not only by mitigating swelling, but also by enhancing CAR T cell efficacy.

Results from monoclonal antibody therapy as a monotherapy and the clinical trial published by Bagley et al. (2024) where CAR T cells were combined with intravenous infusion of anti-PD1 antibodies, were not successful [31, 162, 163]. Since these results are at least partly due to the limited crossing of the BBB by the antibodies after systemic delivery, other approaches are necessary. For instance, since CARs are shown to traffic over the BBB, using CARs that express and excrete immune checkpoint inhibitors will be more effective. Also, other routes of administration of the antibodies need to be tested, because locoregional administration, such as intratumoral infusion, could circumvent the poor trafficking over the BBB by the antibodies. If, in the future, these strategies improve the efficacy of immune checkpoint inhibitors, they can also be tested in combination with CAR T cell therapy for treating pHGG, because immune checkpoint inhibitors such as PD-1 and TIM3 can modulate the TIME towards a pro-inflammatory TIME, which will enhance CAR T cell anti-tumor activity [164].

Novel promising cancer therapeutics, such as ONC201, oncolytic viruses and ultrasound therapy might also be combined with CAR T cell therapy in the future. ONC201 is a novel promising therapy for DMG, which might be combined with CAR T cell therapy in the future [165]. Ma et al. (2021) found that oncolytic viruses expressing IL15/IL15Rα and anti-EGFR CAR NK cells showed synergistic effect in a glioblastoma murine model [166]. They also found that the oncolytic viruses reduced the exhaustion of the NK cells. This strategy could be applied to CAR-based therapies in pHGGs to prevent CAR cell exhaustion, which is likely to occur in the immunosuppressive TIME of pHGG. Recently, a conference abstract was published in which they reported that ultrasound therapy in a glioblastoma murine model can induce trafficking of T cells towards the tumor and in parallel, induce myeloid cells to traffic to the lymph nodes [167]. This novel strategy is promising for treating pHGG in a combination therapy with CAR T cell therapy, because ultrasound therapy could induce CAR T cell infiltration into the pHGG.

### 3.7. Challenges

Despite promising results from preclinical and clinical studies with CAR-based strategies in HGG, curative results remain absent. A rationale for this lack of clinical efficiency may be that although the CAR therapy can achieve reduction of the glioma size, the tumor is not eliminated completely, resulting in tumor recurrence or treatment resistance. Besides promising developments and prospects on CAR based therapy for pHGG, there are also some challenges to overcome. The non-inflammatory TIME in DMG and the immunosuppressive TIME in other pHGG, with low numbers of infiltrating T cells, can be a hurdle for CAR T cell therapy [133]. However, recent studies are focusing on other CAR cell types besides T cells, such as NK or myeloid cells, which are thought to infiltrate gliomas more effectively. In addition, combination therapies or novel CAR T cell designs can also overcome the negative effect of the TIME, by modulating the TIME towards a more pro-inflammatory TIME.

In addition, manufacturing of the CAR T cells might also be problematic, given the time it takes and corresponding costs. For example, for CAR T cell production, clean rooms and CAR T cell facilities are needed. In addition, the process of CAR T cell therapy, including lymphodepletion and CAR T cell manufacturing, may be a limiting factor for pHGG patients, whose life expectancy is often only a few months with rapid tumor progression. Another challenge in pHGG is the lack of available TAAs, since most research only focused on adult glioblastoma. In addition, most TAAs that are found, are heterogeneously expressed in pHGG, which are notorious for their heterogeneity. This heterogeneous target signature can result in antigen escape and may require novel multi-targeted strategies.

Lastly, there are some limitations to this review, including the fact that not all ongoing and completed clinical trials are well documented on websites such as clinical trials.gov. Therefore, the authors cannot guarantee that the overview of clinical trials is complete. In this review, we did not include LGGs, because the LGG research field is distinct from the HGG research, and also because there is limited research describing CAR-based therapeutic targets for LGG, there is no need for a review of the literature yet. Despite the challenges mentioned, this review gives an overview of the promising targets for each of the HGG entities, future research could focus on these targets, maybe even combine them in a multi-target approach.

## Materials and Methods

We performed a systematic literature search using Scopus with query: “TITLE-ABS ((glioma OR (glia AND tumor) OR glioblastoma OR GBM OR astrocytoma OR oligodendroglioma OR (angiocentric AND glioma) OR (diffuse AND midline AND glioma) OR DMG OR DIPG OR ependymoma OR (ependymal AND tumor)) AND (CAR OR (chimeric AND antigen AND receptor) OR (T AND cell AND target AND (immuno OR immune) AND therapy)) AND (target)) AND EXCLUDE(DOCTYPE, “re”)”. This search resulted in 300 articles and book chapters, and we selected articles based on relevancy by reading the title and abstract.

Subsequently, we read and analyzed the papers, as well as relevant papers from the bibliographies. We added relevant papers by hand to have a complete review. In addition, we added relevant preprints and conference abstracts. We also performed a search in clinicaltrails.gov accessed on 15 January 2024 with query “glioma OR (brain AND tumor)” for condition/disease and “CAR” for intervention/treatment, which showed a total of 61 registered clinical trials. We remained with 48 trials, after elimination of withdrawn (5) and irrelevant (8) trials. We included 11 trials that were mentioned in articles of the literature search.

## Conclusions

This review provides an overview of the targets used in clinical trials evaluating CAR-based therapy in pHGG. Immunotherapy research for the treatment of pHGG is still in a preliminary phase, however, alternative CAR-based strategies are under eager investigation by the scientific community. Nevertheless, more preclinical and clinical research is desperately needed to overcome challenges, such as the immunosuppressive TIME, heterogeneous expression of TAA and treatment-related toxicity. Further research should focus on optimization of the CAR design, multi-targeting approach, using CAR NK or myeloid cells and combination therapies for improved CAR-based therapy for pHGG. Finally we conclude that, since the TIME, TAA expression and toxicity vary across pHGG entities, future research should focus on designing entity-specific CAR-based treatments.

## Data Availability

All data produced in the present work are contained in the manuscript

---

**Supplementary table 1.**
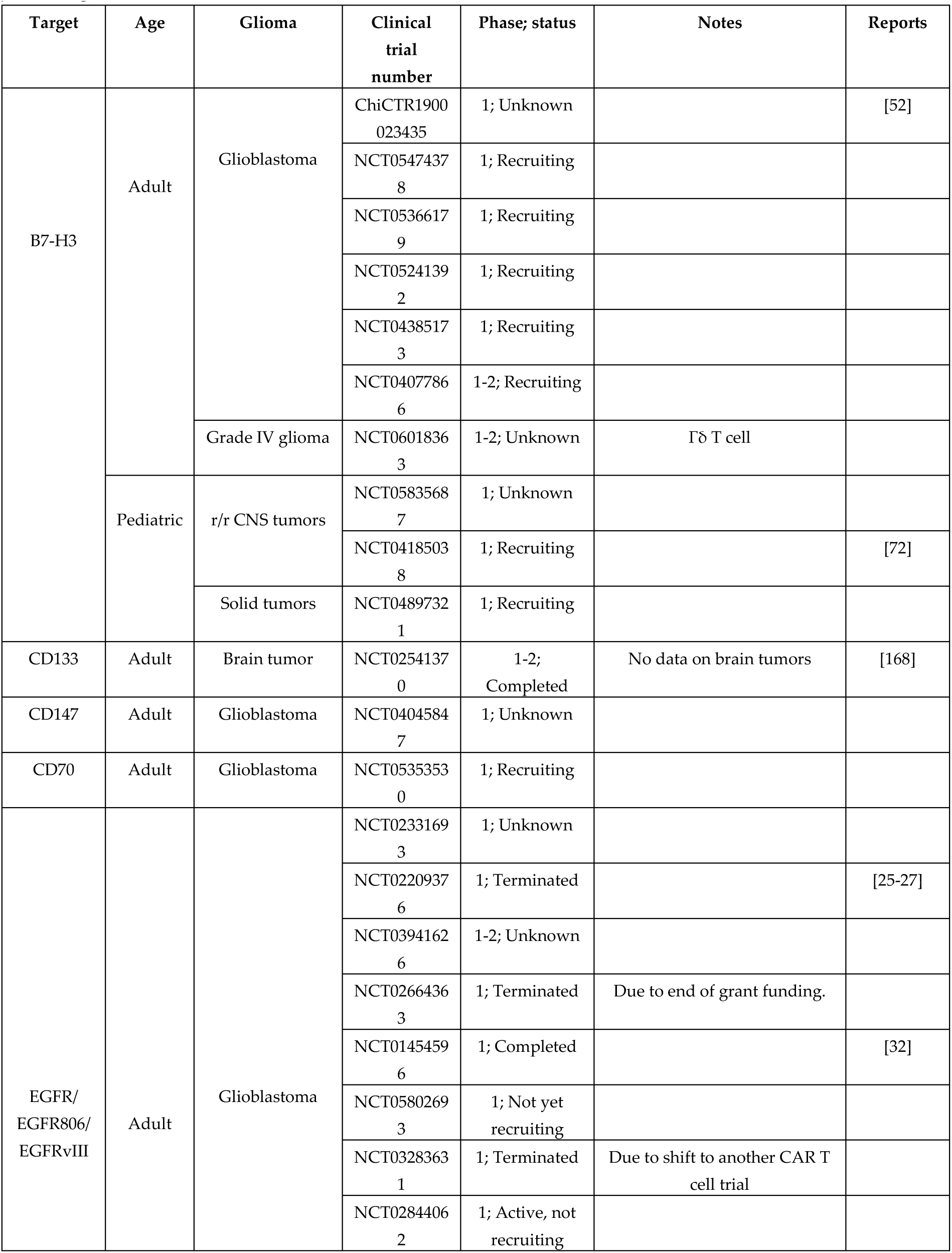
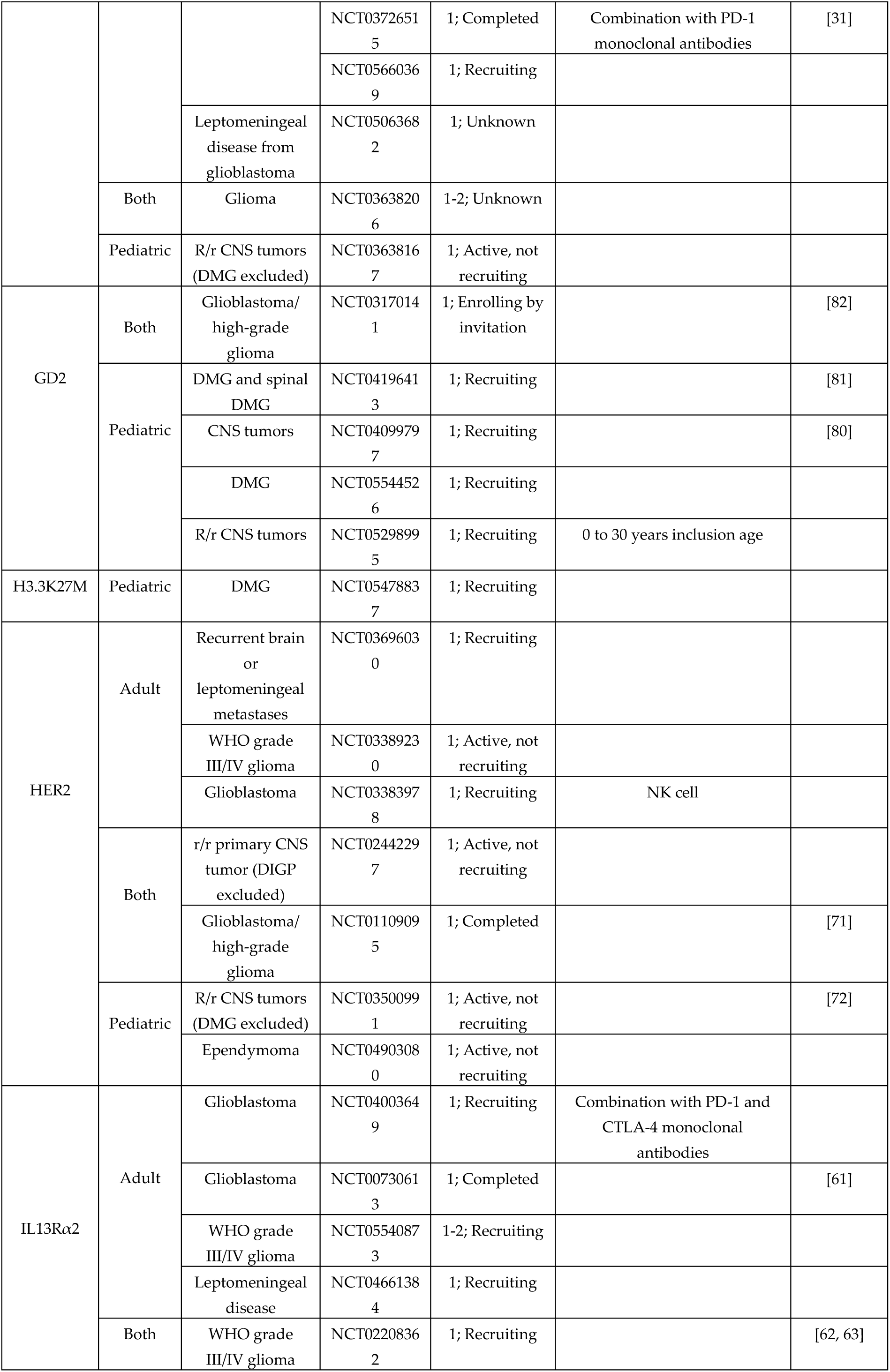
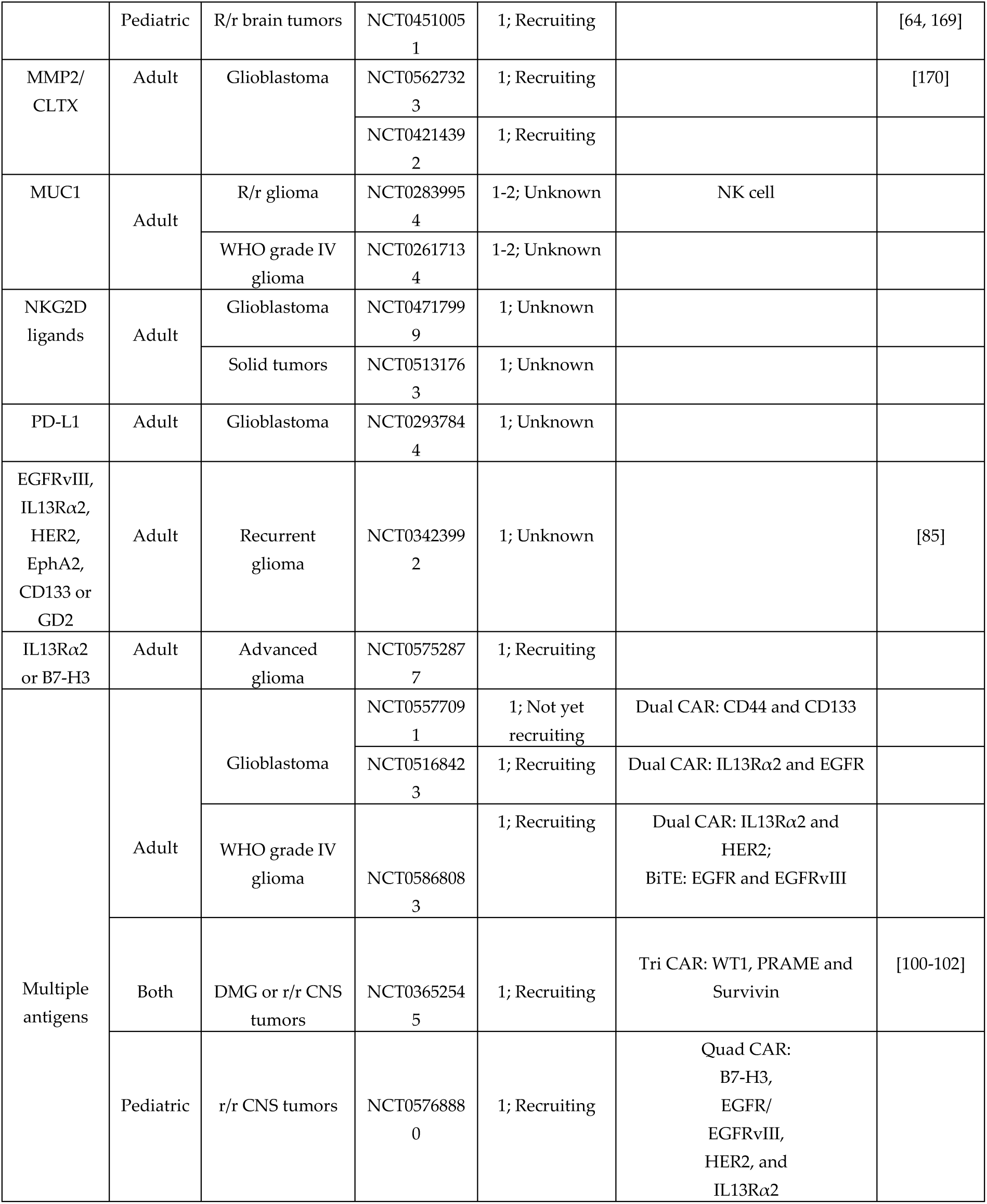
An overview of the current completed and ongoing clinical trials on CAR-based therapy for adult and pediatric glioma.

**Supplementary table 2.**
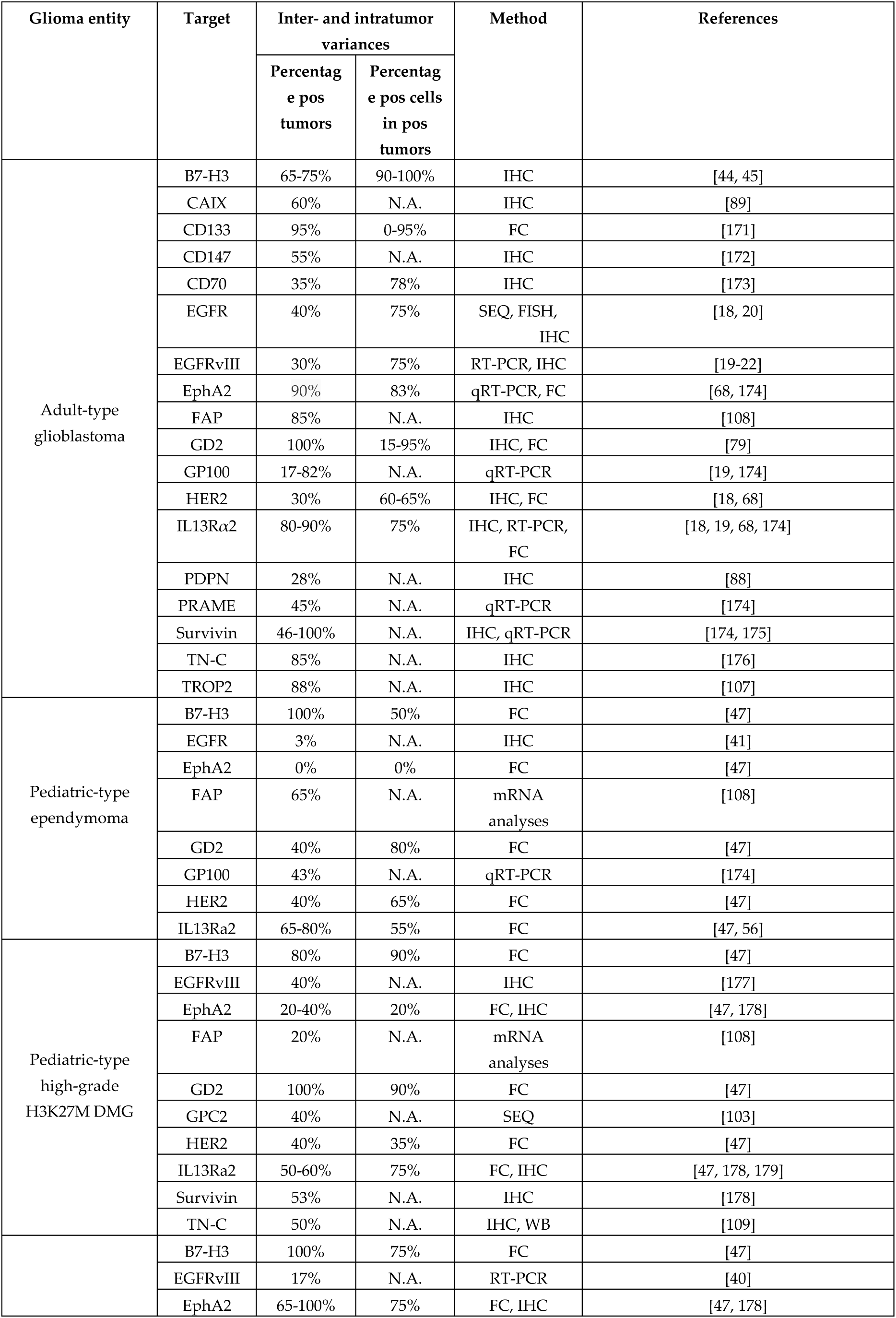
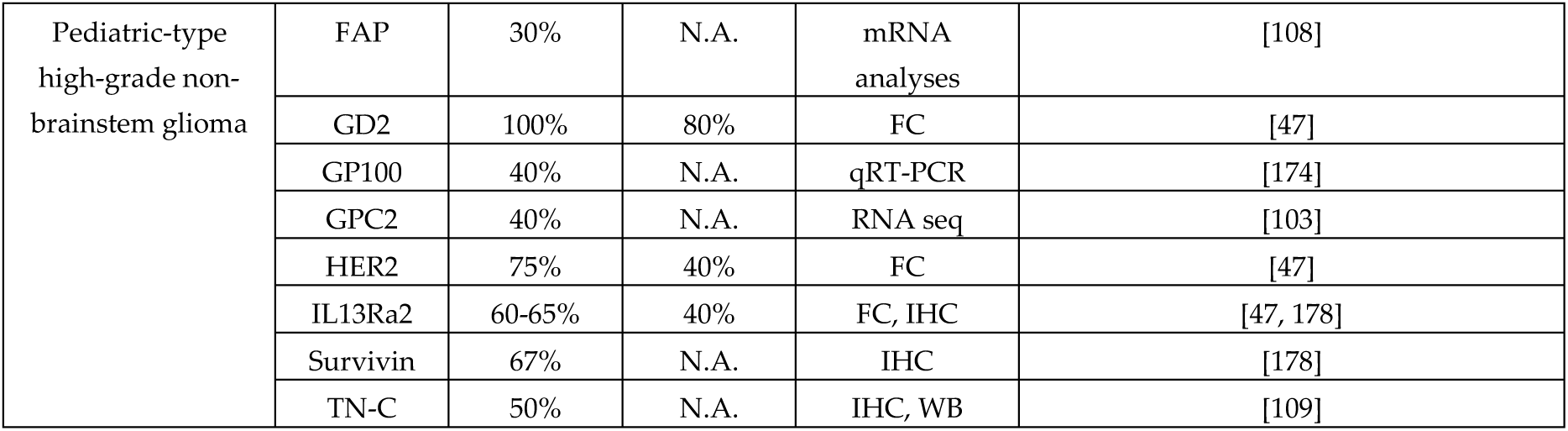
An overview of the CAR-based therapeutic targets and their expression for adult and pediatric glioma. IHC: Immunohistochemistry; SEQ: sequencing; FISH: fluorescence in situ hybridization; qRT-PCR: Quantitative reverse transcriptase polymerase chain reaction; FC: flow cytometry; WB: Western blot

